# Population-level Impact of HPV Vaccination On the Incidence of Genital Warts in Sweden

**DOI:** 10.1101/2024.09.19.24313952

**Authors:** Ana Martina Astorga Alsina, Eva Herweijer, Jiayao Lei

## Abstract

**Background:** Sweden introduced HPV vaccination in 2006, administered through opportunistic, subsidized, catch-up and school-based programs. Notably, genital warts (GW) are the first observable clinical outcome following infection by HPV-6/11, targeted by vaccination. We aim to gain knowledge of the population incidence of GW in Sweden and evaluate its change throughout vaccination programs.

**Methods:** This ecological study used Swedish registers to obtain national population data and cases of genital warts from 2006-2018 in men and women aged 15-44. We used Poisson models to evaluate GW incidence change after vaccination in reference to a pre-vaccination period, stratified by age and sex. As well as, to estimate incidence change of GW in birth cohorts eligible for different vaccination programs compared to a pre-vaccination cohort. Finally, we estimated GW cases averted in each vaccinated cohort.

**Results:** The incidence of GW decreased during periods following HPV vaccination. In 2016-2018, over a decade after vaccination availability, incidence decreased by 89% (95% CI 83-93), 73% (95% CI 71-75), 50% (95% CI 43-56) and 20% (95% CI 10-28) in women aged 15-19, 20-24, 25-29 and 30-34, respectively. A similar reduction was observable in men, although of lesser magnitude. We estimated 18,890 and 12,343 GW cases averted among vaccinated cohorts of women and men, respectively.

**Conclusions:** We report on population-level decreases of GW incidence in women and men following increased vaccination coverage. Cohorts eligible for school-based vaccination recorded the largest decrease in GW incidence in Sweden to date. For the period under study, decreases among men could be attributed to herd effects.

## Introduction

In 2020, the World Health Organisation (WHO) proposed a global strategy to eliminate cervical cancer as a public health issue, aiming to achieve 90% vaccination coverage against human papillomavirus (HPV) in girls by age 15 [1]. In Sweden, immunisation against HPV was first offered in October 2006, with almost exclusive administration of the quadrivalent vaccine (i.e. covering HPV-6/11/16/18) until 2019 [2-4]. Initially, vaccination was opportunistic and required an out-of-pocket fee but was subsidised from May 2007 (supplementary Figure S1 & Table S1) [2]. National coverage remained low until vaccination delivery changed to a structured school-based program in 2012, accompanied with a catch-up program, and no out-of-pocket fee [3-4]. Since then, coverage of at least one dose increased to 80% and 60% for the school-based program cohorts and catch-up program cohorts, respectively [6]. Later, in 2016, another catch-up opportunity came within the school-based program for those under 18 [4]. Males were mostly excluded from national vaccination programs until the extension of school-based vaccination in autumn 2020 [5].

Monitoring the impact of changing HPV vaccination coverage on cervical cancer incidence can be challenging due to the prolonged latency period. Clinical outcomes of HPV that precede cancer, such as cervical lesions or genital warts (GW), are thus useful to monitor potential impact at an earlier stage. 90% of GW cases are associated with HPV-6/11, covered by quadrivalent and nonavalent vaccines [7]. They may develop as early as 2-3 months from the point of infection and are widely reported by healthcare registers in Sweden [7]. As a result, they serve as the first observable disease measure to assess early effectiveness of HPV vaccines [8,9].

Several countries have reported reductions of GW incidence following HPV vaccination program implementation [10–13]. However, in Sweden, GW are not included in the mandatory surveillance of infectious diseases, and incidence has only been reported up until 2012 [3,6,14,15]. Thus, to date, the school-based vaccination program change has not been evaluated for its population impact on GW.

We aim to examine the incidence trend of GW from 2006 to 2018 among 15-44 year old men and women in Sweden, in relation to subsequent national HPV vaccination programs. The inclusion of men allows for exploration of possible protection via herd-effect since their coverage during the study period is virtually zero. We also estimate cases of GW averted among birth cohorts eligible for Sweden’s sequential HPV vaccination programs.

## Methods

### Study Population & Data Collection

Our study followed an ecological time-trend design, where GW time-varying disease rates were retrospectively compared in a population geographically limited to Sweden. The study’s target population consists of men and women aged 15 to 44, registered in Sweden between 2006 to 2018.

We collected aggregated Swedish population data from the Total Population Register (TPR), maintained and reported by Statistics Sweden. The TPR holds registration of all individuals permanently residing in Sweden since 1968 [16]. We also obtained counts of GW cases through healthcare register data from the National Patient Register (NPR) using the ICD-10 code denoting an anogenital wart diagnosis (A63.0) [17], and from the Prescribed Drug Register (PDR) using ATC codes for podophyllotoxin (D06BB04) and imiquimod (D06BB10) [18]. Non-pharmacological treatments used in persistent cases after first-line pharmacological treatment were excluded.

### Vaccination Coverage

We defined exposure categorically based on HPV vaccination coverage at the population-level, in relation to Sweden′s sequential vaccination programs. Periods of pre- and post-vaccination were defined via coverage reported by the Public Health Agency of Sweden, which refers to all those who have received at least one dose of a HPV vaccine [5]. We defined the pre-vaccination period as 2006-2007 because vaccination was introduced late in 2006 and coverage remained close to zero during this period (supplementary Figure S1). The post-vaccination periods include 2008-2011, 2012-2015 and 2016-2018, which registered sequentially increased coverage (i.e. specific to each birth cohort), of up to 34%, 55% and 84%, respectively [5].

Subsequently, these categorical classifications were made based on birth cohort eligibility for each national program (supplementary Table S1 and Figure S2). Pre-vaccination birth cohorts include 1961-1985, who were not targeted by any national vaccination programs, and 1986-1988, who may have been vaccinated opportunistically but without subsidy (i.e. coverage does not exceed 10%) [5]. Meanwhile, post-vaccination birth cohorts include 1989-1992, who were part of the subsidised vaccination program, 1993-1998, who were eligible for the subsidised vaccination program and the 2012 catch-up program, as well as 1999-2003, who participated in the school-based vaccination program.

### Genital Wart Cases

Cases of GW are considered a sufficiently specific outcome that allows measurement of vaccination effects as 90% of cases are ascribed to HPV-6/11, which are targeted by the quadrivalent vaccine in use during the study period [7, 19]. We included both first-occurring and subsequent cases of GW from any single individual, since no differentiation was made between these. A new case was included in the absence of any GW-related clinical or pharmaceutical registers in the 6 months prior. We pooled counts of GW cases, aggregated by sex, age-at-diagnosis, period of diagnosis and birth cohort.

### Statistical Analyses

We report descriptive plots, a period analysis and a cohort analysis performed using Stata (version 18). We first merged population and case data (see supplementary Section S1:Methods) [20]. Then, our descriptive analyses involved crude estimates for incidence rate (IR) of GW, derived from the number of GW cases divided by the corresponding person-years at risk. We plotted age-specific incidence by various calendar periods, and period-specific incidence by age groups [21]. These were illustrated for women and men respectively. In the period analysis that followed, we estimated incidence rate ratios (IRR) and 95% confidence intervals (CI) for post-vaccination periods using the pre-vaccination period (i.e. 2006-2007) as reference. To do so, we employed Poisson regression models with a categorical period variable as the predictor and robust standard errors, stratifying by each age and sex category.

After this, we conducted a cohort analysis to estimate IRR with 95% CIs for each birth cohort group eligible for subsequent vaccination programs using the youngest pre-vaccination cohort, 1986-1988, as reference. We used a Poisson regression model with robust standard errors, and a categorical cohort variable as predictor. As covariates, we included age and period as restricted cubic splines with 6 knots and 3 knots, respectively. We then estimated the amount of GW cases averted with 95% CIs for each post-vaccination birth cohort group. This was done by first estimating the number of GW cases in each vaccinated cohort using each cohort’s different estimated effects and size. Then, we compared this with the corresponding expected number of GW cases when the cohort effect was forced to be the same as in the referent pre-vaccination cohort using the margin command in Stata [22]. The difference between estimated and expected GW cases was considered as the number of GW cases averted through HPV vaccination.

Finally, we performed sensitivity analyses to evaluate robustness in our estimates. In the period analysis, we explored the sensitivity of our estimates to an adjustment for seasonal effects. This involved running the same model for each age and sex category with the inclusion of a categorical season variable as a covariate. In the cohort analysis, a negative binomial model was included to compare model sensitivity using the same predictors and covariates as the main model. Using this model, we again estimated IRR and subsequently, the number of cases averted.

## Results

Our study included a total of 184,006 incident cases of GWs among 15-44 year old Swedish residents from 2006 to 2018, including 80,296 cases detected in women and 103,710 in men (Table 1). The study population accumulated a total of 23.64 million person-years from women and 24.83 million from men throughout the study period.

**Table 1:** Sex-specific genital warts cases and person-time per categories of age, period, and cohort.

|  | Men |  |  |  | Women |  |  |  |
| --- | --- | --- | --- | --- | --- | --- | --- | --- |
|  | Genital Warts |  | Person-Years |  | Genital Warts |  | Person-Years |  |
|  | n=103,710<br>Cases | % | n=24.83 million<br>Millions | % | n=80,296<br>Cases | % | n=23.64 million<br>Millions | % |
| **Age** |  |  |  |  |  |  |  |  |
| 15-19 | 6,198 | 5.98 | 3.9 | 15.66 | 13,655 | 17.01 | 3.6 | 15.34 |
| 20-24 | 31,570 | 30.44 | 4.2 | 16.80 | 25,955 | 32.32 | 3.9 | 16.68 |
| 25-29 | 30,469 | 29.38 | 4.2 | 17.08 | 17,394 | 21.66 | 4.0 | 17.06 |
| 30-34 | 17,717 | 17.08 | 4.1 | 16.49 | 10,084 | 12.56 | 3.9 | 16.51 |
| 35-39 | 10,588 | 10.21 | 4.1 | 16.65 | 6,990 | 8.71 | 4.0 | 16.83 |
| 40-44 | 7,168 | 6.91 | 4.3 | 17.33 | 6,218 | 7.74 | 4.2 | 17.58 |
| **Calendar Year** |  |  |  |  |  |  |  |  |
| 2006-2007 | 16,993 | 16.39 | 3.7 | 14.97 | 16,165 | 20.13 | 3.6 | 15.05 |
| 2008-2011 | 37,862 | 36.51 | 7.5 | 30.39 | 31,319 | 39.00 | 7.2 | 30.55 |
| 2012-2015 | 31,121 | 30.01 | 7.6 | 30.71 | 21,334 | 26.57 | 7.3 | 30.75 |
| 2016-2018 | 17,734 | 17.10 | 5.9 | 23.93 | 11,478 | 14.29 | 5.6 | 23.65 |
| **Birth Cohort** |  |  |  |  |  |  |  |  |
| 1961-1985 | 60,467 | 58.30 | 15.2 | 61.11 | 39,396 | 49.06 | 14.5 | 61.54 |
| 1986-1988 | 18,911 | 18.23 | 2.6 | 10.53 | 16,072 | 20.02 | 2.5 | 10.59 |
| 1989-1992 | 18,395 | 17.74 | 3.5 | 14.14 | 17,932 | 22.33 | 3.3 | 14.05 |
| 1993-1998 | 5,786 | 5.58 | 2.8 | 11.35 | 6,688 | 8.33 | 2.6 | 11.04 |
| 1999-2003 | 151 | 0.15 | 0.7 | 2.87 | 208 | 0.26 | 0.7 | 2.78 |

In the descriptive figures, we noticed a difference in age trends of GW incidence between women and men where the maximum IR among women in 2006-2007 is seen at age 21 (1178 cases per 100,000 person-years), while among men at 23 (1167 cases per 100,000) (Figure 1). Besides this, however, crude curves for GW cases by age-at-diagnosis appeared mostly similar among women and men, with 71% of cases occurring before 30 among women and 66% among men (Table 1).

**Figure 1:**
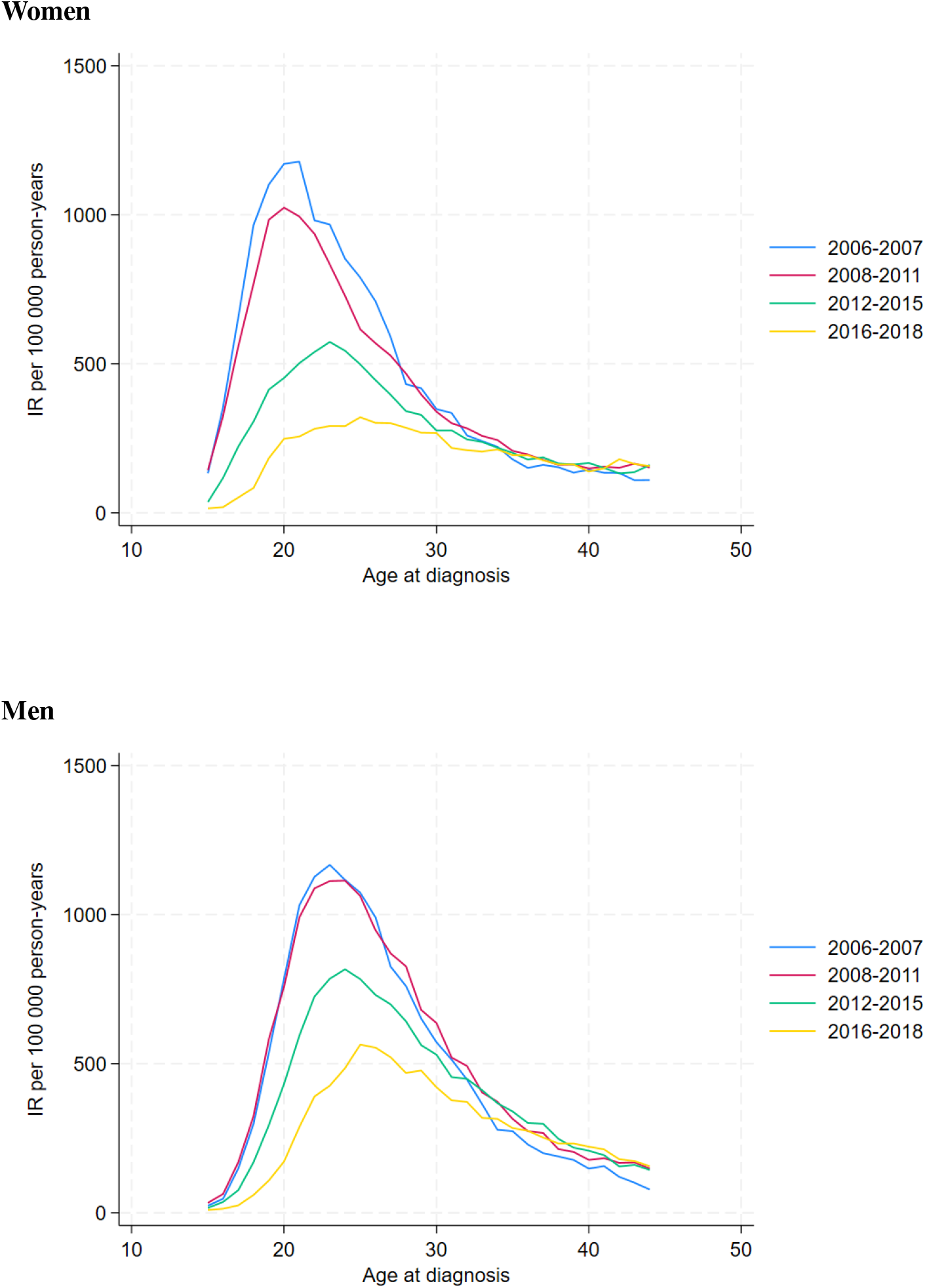
Age-trend of genital warts incidence rates among women and men, stratified by period

Then, when inspecting crude IR over calendar time visually, we observed a declining pattern of varying magnitude among those aged 15-19, 20-24 and 25-29 (Figure 2). This was also reflected in the flattening of the age curve across period strata (Figure 1). Among women, the decline appeared to begin from 2007/2008 while in men it appeared later around 2010 (Figure 2).

**Figure 2:**
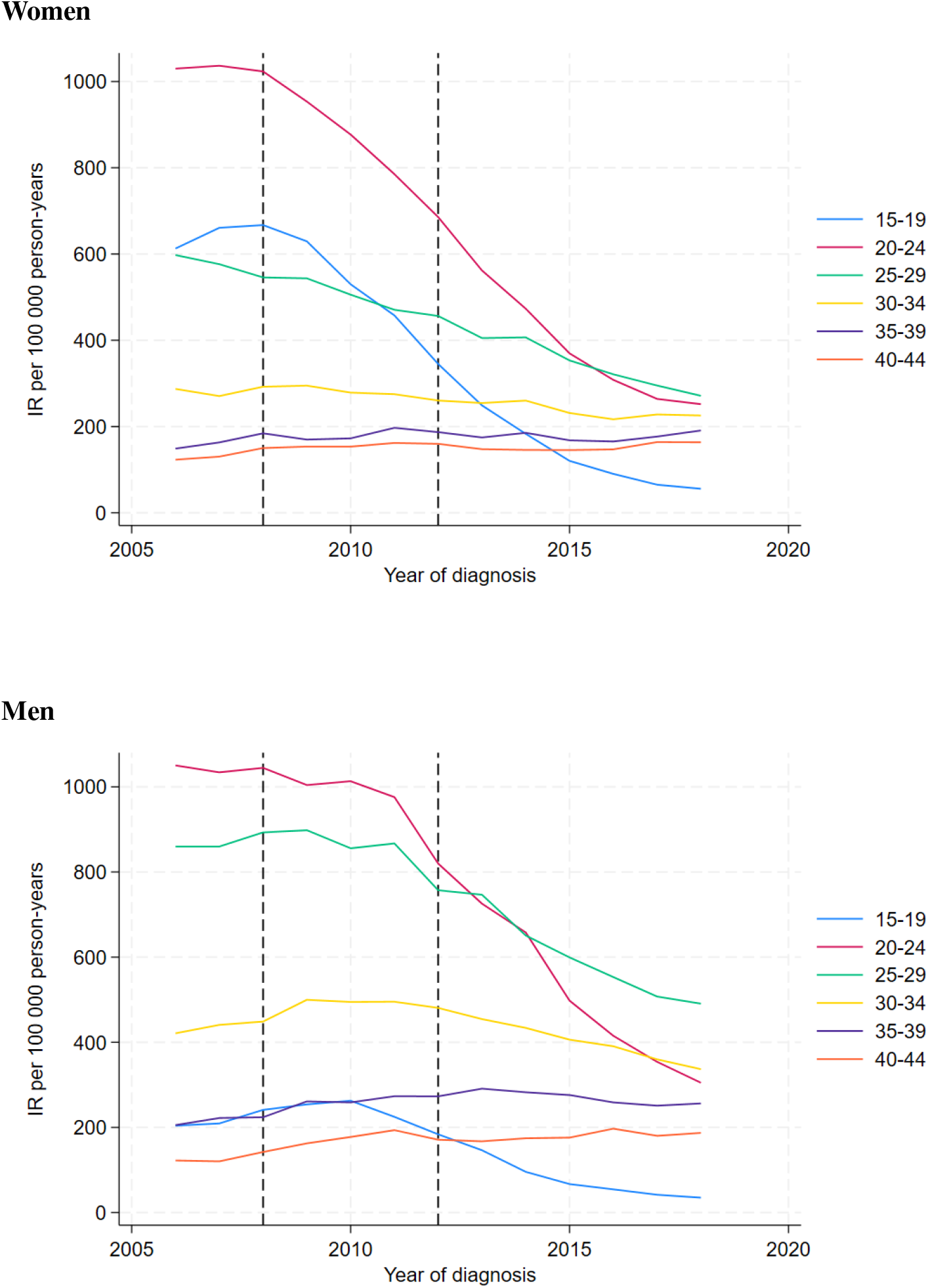
Incidence rates of genital warts among women and men between 2006 and 2018, stratified by age group. Dashed vertical line at 2008, denoting start of post-vaccination period, and at 2012, denoting start of school-based vaccination period

Similarly, through the Poisson model of the period analysis, we estimated the largest and earliest decreases in IR of GW among women aged below 30. This was for each consecutive period in reference to the pre-vaccination period (i.e. 2006-2007) (Figure 3, supplementary Table S2). Among this group, we could clearly observe a pattern of increased IR reductions in later time periods. Among 15-19 year olds, for example, IRR estimates showed decreases of 10% (95% CI (−22)-33), 64% (95% CI 50-75) and 89% (95% CI 83-93) for 2008-2011, 2012-2015 and 2016-2018, respectively. Besides this, we could also observe a pattern of decreased reductions with increased age. This difference is observed in the comparably smaller estimates among 20-24 year olds of 12% (95% CI 5-19), 49% (95% CI 44-54) and 73% (95% CI 71-75) for 2008-2011, 2012-2015 and 2016-2018, respectively. Meanwhile among 25-29 year olds we estimated this decrease at 12% (95% CI 0-23), 31% (95% CI 21-40) and 50% (95% CI 43-56) for the same time periods, respectively.

**Figure 3:**
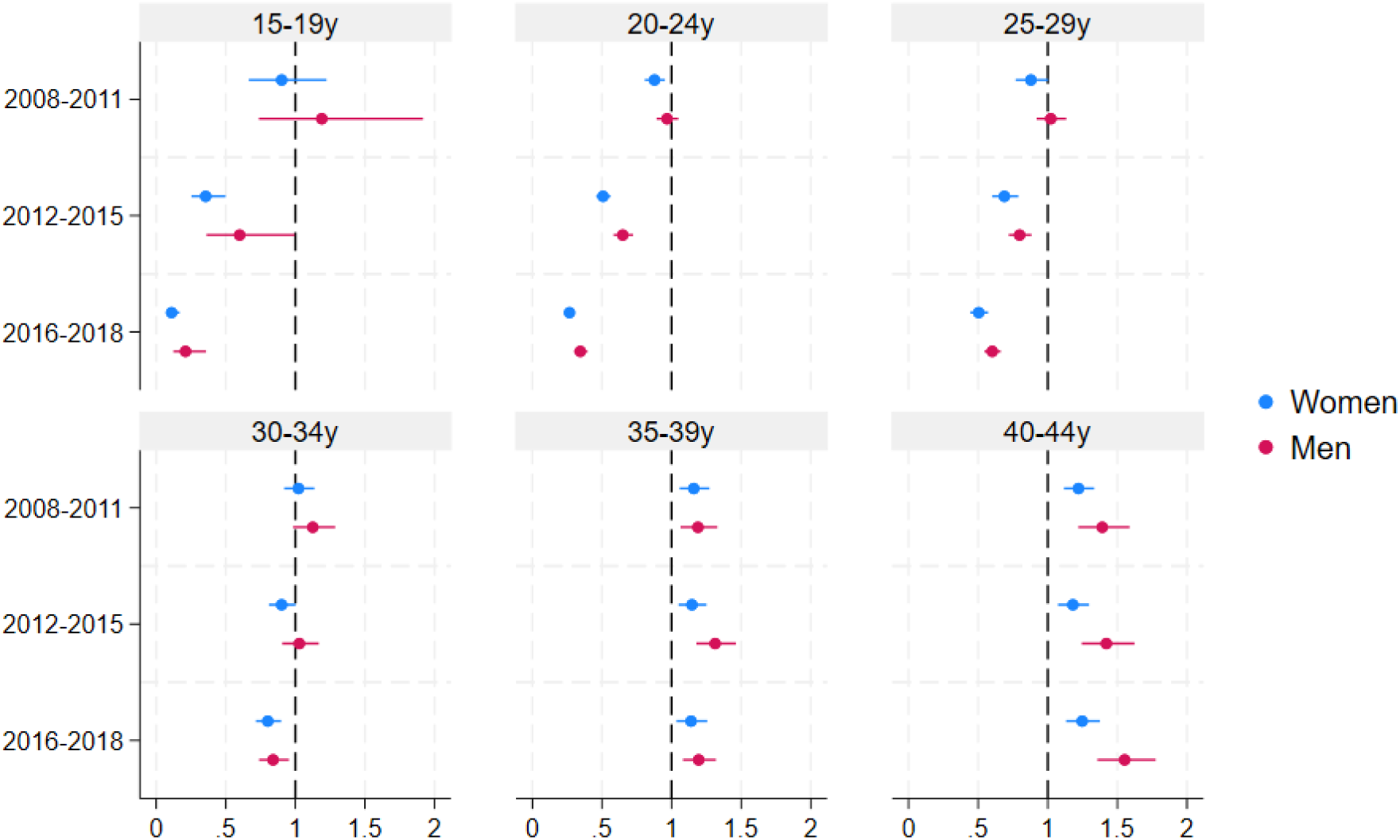
Age-specific incidence rate ratio of genital warts cases for post-vaccination time periods in reference to pre-vaccination (i.e. 2006-2007), stratified by sex

Among women aged above 30, on the other hand, we observed stable IR over time compared to the reference pre-vaccination period (Figure 3, supplementary Table S2). Unlike women aged below 30, we generally didn’t observe a consistent pattern of increasing IR reductions for later time periods in this age group. This could only be observed in women aged 30-34, whose estimates in 2012-2015 begin to show a decrease in IR of 10% (95% CI 0-19), and later 20% (95% CI 10-28) in 2016-2018. Estimates of women aged 40-44 in 2016-2018 showed the largest increases in GW IR at any period of 25% (95% CI 13-38).

When it comes to men, we observed comparable patterns to those among women, including increased reductions of IR over time and at younger ages. Although, estimates of IR decreases were lower than their female counterparts in those under 30, with the largest estimated decreases among those aged 15-19 in 2012-2015 and 2016-2018 being 40% (95% CI 1-64) and 79% (95% CI 64-88), respectively (Figure 3, supplementary Table S2). Meanwhile estimates of IR increase among men over 30 were higher compared to women, reaching 55% (95% CI 36-78) among 40-44 year olds in 2016-2018. We also generally observed wider CIs among men compared to women.

### Comparison Of Birth Cohorts

Cohort analyses showed significant changes in incidence of GW in post-vaccination birth cohorts compared to the reference birth cohort, 1986-1988 (Table 2). With the Poisson model, we estimated a decrease in incidence of 23% (95% CI 18-27), 62% (95% CI 58-65) and 93% (95% CI 90-95) in birth cohorts of women eligible for subsidised, catch-up and school-based vaccination, respectively. Meanwhile, in male birth cohorts that corresponded to cohorts of women eligible for vaccination, we saw similar decreases in risk, of 19% (95% CI15-22), 56% (95% CI 52-59) and 85% (95% CI 81-88), respectively.

**Table 2:**
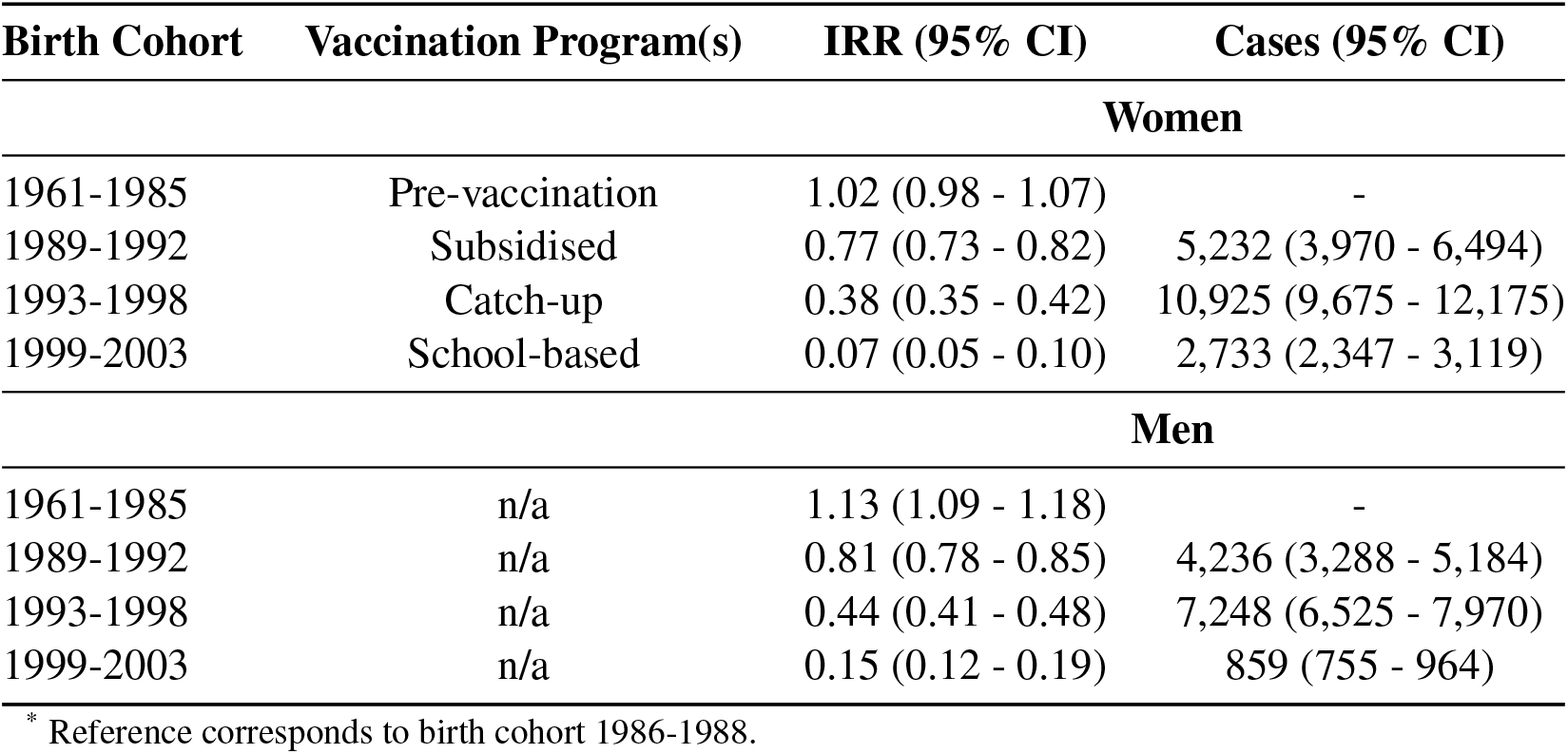
Incidence Rate Ratio (IRR) of genital warts and estimated number of cases averted per birth cohort.

| Birth Cohort | Vaccination Program(s) | IRR (95% CI) | Cases (95% CI) |
| --- | --- | --- | --- |
| **Women** |  |  |  |
| 1961-1985 | Pre-vaccination | 1.02 (0.98 - 1.07) | - |
| 1989-1992 | Subsidised | 0.77 (0.73 - 0.82) | 5,232 (3,970 - 6,494) |
| 1993-1998 | Catch-up | 0.38 (0.35 - 0.42) | 10,925 (9,675 - 12,175) |
| 1999-2003 | School-based | 0.07 (0.05 - 0.10) | 2,733 (2,347 - 3,119) |
| **Men** |  |  |  |
| 1961-1985 | n/a | 1.13 (1.09 - 1.18) | - |
| 1989-1992 | n/a | 0.81 (0.78 - 0.85) | 4,236 (3,288 - 5,184) |
| 1993-1998 | n/a | 0.44 (0.41 - 0.48) | 7,248 (6,525 - 7,970) |
| 1999-2003 | n/a | 0.15 (0.12 - 0.19) | 859 (755 - 964) |

\* Reference corresponds to birth cohort 1986-1988.

Using IR of the reference cohort as a counterfactual scenario allowed us to estimate cases of GW averted in each birth cohort. This way, we estimated 31,233 cases of GW averted in cohorts that were eligible for any structured national HPV vaccination program, compared to what would have been expected if the cohort effect had been the same as the reference pre-vaccination cohort’s. This included 18,890 cases among women and 12,343 cases among men (Table 2). The largest number of cases averted were in cohorts 1993-1998 followed by 1989-1992 and then 1999-2003. This is true in both sexes while the estimated number of cases averted is lower in men, and this difference is larger in younger cohorts.

### Sensitivity Analysis

Sensitivity analyses showed robustness of our estimates to modified assumptions. In the period analysis, our estimates were comparable after adjustment for seasonality (supplementary Table S3): changes in the magnitude of associations were negligible, with those among men aged 15-19 being the largest, while CIs were comparable. In the cohort analysis, model-sensitivity analysis showed robustness to specification error, with the results produced by the negative binomial model being very similar to the main analysis: IRRs estimated from the negative binomial model were generally slightly larger, and consequently led to larger estimates of cases averted (supplementary Table S4), while CIs were comparable.

## Discussion

We demonstrate a decreased incidence of GW after the introduction of HPV vaccination, with a larger decrease over time as vaccination coverage increased. This trend is clearly observed in women below the age of 30, who were eligible for a structured vaccination program. This age group also displays a clear trend of reduced decreases in IR with older age, correlating with a decrease in coverage with increased age. In contrast, the estimated IR for women aged over 30 (i.e. born before 1989) generally remained approximately stable. This population would generally not have had access to subsidised vaccination, and coverage would be vulnerable to disparities influencing opportunistic access. An exception is observed among those aged 30-34 who have a decreased IR after 2012, likely due to some coverage from the opportunistic vaccination period prior to subsidisation While IR trends over the study period observed in men resembled those observed in women, some differences allude to their differential protection. IRR estimates among men were generally more positive, indicating lesser magnitude decreases and higher magnitude increases in incidence. This alludes to a lower protective effect among men, in-line with their general lack of direct protection against HPV. Among those under 30, estimates showed no significant trends until 2012-2015. This indicates a lag in effect in this population compared to women. Men were only included in a national HPV vaccination program in Sweden from autumn 2020. Thus, if these trends were attributed to protection via a vaccination program, it could only be indirect protection via herd effect.

Additionally, we report the lowest estimated IRR in birth cohort group 1999-2003. Despite this, most cases averted corresponded to birth cohort, 1993-1998, who were likely protected via a mix of the opportunistic and catch-up program (supplementary Table S1). In this group, vaccination coverage is relatively high, and they are included in the study throughout peak infection ages, thereby accounting for many cases averted. While coverage is also high for cohorts 1999-2003, the study period only included them before the age of maximum incidence of GW (supplementary Figure S3). Similarly, even though coverage is lower in cohorts born 1989-1992, their inclusion in the study during a wide age range of high incidence of GW resulted in many averted cases estimated.

Overall, our results indicated that GW incidence sees a higher decrease over time and lower decrease with increased age during the study period. This could be attributed to higher coverage and inclusion of younger cohorts, who may have also been vaccinated at an earlier age. We also indicate a possible increased herd effect in men over time. These observations are reliant on GW diagnostics remaining constant during the study period, and do not consider differences in protection from differential vaccine doses. While it is difficult to definitively differentiate between the effect of distinct vaccination programs, the introduction of subsidisation and structured delivery in school-settings likely facilitated convenience for vaccination uptake and resulted in the observable increase in vaccination coverage; consequently, a comparable potential impact on incidence of GW seems plausible.

Our results corroborate with existing reports of GW trends in earlier time periods following HPV vaccination in the Swedish population. This includes a decrease in IR of GW among women aged 15-24 from 2008, and among men from 2010-2012 [6]. However, while previous studies state no change in GW IR for women above 30 years of age, we report decreases in women aged 30-34 from 2012 onwards and in men from 2016 [6]. This finding is ascribed to our study’s prolonged follow-up since vaccine introduction.

Furthermore, our results align with literature from other countries concerning impact of vaccination programs on incidence of GW. Comparable estimates from a Danish population registry study of GW incidence 5 years after vaccine implementation found decreases of 55% among women aged 12-35, and 37% among men aged 12-29 [10]. Similarly, an Australian serial cross-sectional study estimated a 68% reduction in GW diagnoses among women aged above 15 and a 49% reduction among men of the same age during the initial girls-only vaccination period [12]. Much like our estimates, they report smaller decreases of GW incidence with increasing age. A Canadian study evaluating the first cohorts eligible for school-based vaccination estimated a GW incidence decline of 72% in women 16-18 years of age and 51% in men of the same age [11]. Meanwhile, summary estimates from a meta-analysis indicated faster and more significant decreases of GW in women aged 15-24 in high coverage contexts (>50%), reinforcing the relationship between GW IR change and the coverage achieved by vaccination programs [13]. Among women aged 25-29 and men aged 15-24, their estimates were only significant in high coverage settings where indirect protection may have had an increased effect. This aligns with our estimates. Meanwhile, among men aged 25-29, the meta-analysis estimated significant effects in high coverage contexts over 5 years post-vaccination like our estimates for the same group, 6 years post-vaccination [13].

There are some limitations in our study. Firstly, GW cases were obtained only for men who are first-degree relatives of the female population, resulting in an underestimation of the true IR when using the person-time of the total population. However, given that our study covers an estimated 96% of the male population in Sweden, we expect this bias to be small. We also cannot account for any HPV vaccinations received before immigrating to Sweden, which may result in an underestimation of coverage. Depending on the overall background risk of GW among immigrants, this could contribute to an over-or underestimation of the impact on GW. Furthermore, some GW cases may be erroneously classified as such through the prescription of podophyllotoxin and imiquimod for other conditions. The most common of these conditions, namely superficial basal cell carcinoma and actinic keratoses, are mostly associated with ages 40 or above [23], minimising bias within the age range of interest. Molluscum contagosium, however, is often found in young, sexually active adults and could lead to an overestimate of GW incidence in our population [24, 25]. This could introduce bias if cases of molluscum contagosium were differential over the study period. Similarly, lack of health-seeking for GW could lead to an underestimation of cases, although this is not expected to be differential over time. Finally, regression of GW cases before identification by either NPR or PDR could contribute to an underestimation of cases. However, GW may often recur and presumably be identified in the national healthcare registers eventually [7].

Furthermore, the ecological time-trend design is prone to ecological fallacy and confounding from time-varying factors. Crucially, our interpretations on impact of vaccination programs are limited to the group-level rather than on any individual. Meanwhile, confounding factors including changing sexual or healthcare-seeking behavior, may also relate to components of the vaccination program that are not vaccination itself, such as increased public campaigns or education of school-personnel. This confounding effect can be partially explored by comparing other STI trends as negative controls.

Nonetheless, we benefit from a series of important strengths. GW cases from the nationwide registers were obtained with near complete coverage of men and women registered in Sweden between 2006-2018. These registers constitute valid and reliable sources for the outcome selected that allow for high statistical power and minimised risk of selection bias. Meanwhile the ecological design allowed for estimates on the total impact of the HPV vaccination program, including both direct and indirect protective effects of the HPV vaccine. This allowed us to report on highly relevant measures at the population level, with significant relevance to the surveillance of vaccination program performance.

Our study provides national-level estimates of the incidence of GW in Sweden 12 years after introduction of HPV vaccination, highlighting trends related to different vaccination delivery approaches. These findings should be generalisable to most settings with similarly structured vaccination programs, achieving comparable coverage rates. Our results fall in-line with recommendations from the WHO and the European Centre for Disease Prevention and Control, which advocate for the increased effectiveness of school-based HPV vaccination at younger ages. By demonstrating the differential population-level impact of various vaccination programs, our findings are crucial for informing policy- and decision-makers on optimizing national immunization programs. Continuous monitoring of the programs and evaluation of GW incidence trends are essential, possibly exploring the potential for eliminating GW through HPV vaccination.

In conclusion, we demonstrate significant decreases of GWs among women under 30 and, more recently, a comparably smaller but sizeable decrease among men through possible herd effect. Cohorts eligible for subsidised national HPV vaccination programs are associated with significantly lower incidence of GW, especially those vaccinated through the school-based program. Ultimately, underscoring the importance of structured and targeted vaccination strategies that minimise cost barriers and maximise convenience, to reduce HPV-related disease outcomes.

## Supporting information

All Supplementary Material

## Data Availability

All data produced in the present study are available upon reasonable request to the authors.

## Ethical considerations

This study was approved by the regional ethical review board in Stockholm (Dnr. 02-556, 2011/921-32, and 2023-06456-02), and concluded that informed consent from the study subjects was not required.

## Funding

This work was supported by the Swedish Research Council (Vetenskapsrådet) (No. 2023-01809), the Swedish Research Council for Health, Working Life and Welfare (FORTE) (No. 2023-01221), and Karolinska Institutet Strategic Research Area in Epidemiology and Biostatistics (JL). The funders have no role in considering the study or in the collection, analysis, interpretation of data, writing of the report, or decision to submit the article for publication.

## Acknowledgements

We would like to acknowledge Prof. Pär Sparen for database establishment and valuable discussion, Pouran Almstedt for data management, as well as Alexander Ploner for consistent statistical support.

## Conflicts of Interest

None to declare.

## Statement of Data Availability

Data not publicly available.

## Notes

### Competing Interest Statement

The authors have declared no competing interest.

### Funding Statement

This work was supported by the Swedish Research Council (Vetenskapsradet) (No. 2023-01809), the Swedish Research Council for Health, Working Life and Welfare (FORTE) (No. 2023-01221), and Karolinska Institutet Strategic Research Area in Epidemiology and Biostatistics (JL). The funders have no role in considering the study or in the collection, analysis, interpretation of data, writing of the report, or decision to submit the article for publication.

