## Supplementary material for "Population-level Impact of HPV Vaccination On the Incidence of Genital Warts in Sweden": All Supplementary Material

September 19, 2024

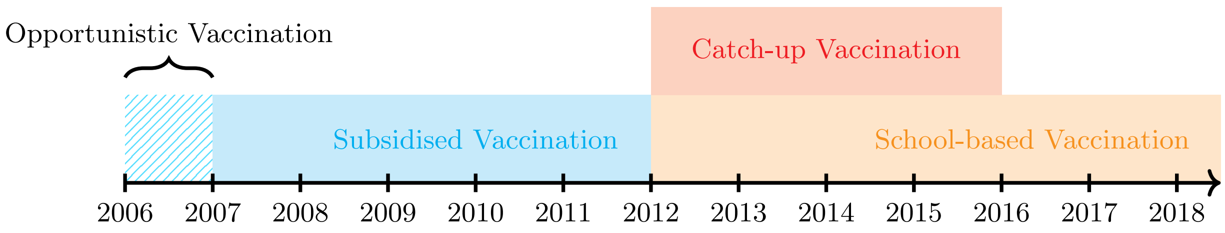


Figure S1: *Active vaccination programs during study period*

Table S1: Populations eligible for each Swedish Vaccination Program

| Vaccination Program | Implementation | Target Age | Sex Targeted | Cohorts Included |
|---|---|---|---|---|
| **Subsidised** | 2007* - 2012 | 13 - 17 | Females | 1989 - 1998 |
| **Catch-up** | 2012 - 2015 | 13 - 18 | Females | 1993 - 1998 |
| **School-based** | 2012 - 2020 | 11 - 13 | Females | 1999 - 2007 |
| | 2016 - present | - 18 | Males** & Females | 1998 - |
| | 2020 - present | 11 - 13 | Males & Females | 2007 - present |

* HPV vaccine was available opportunistically at own cost from October 2006 to May 2007

** Only included in catch-up since 2020

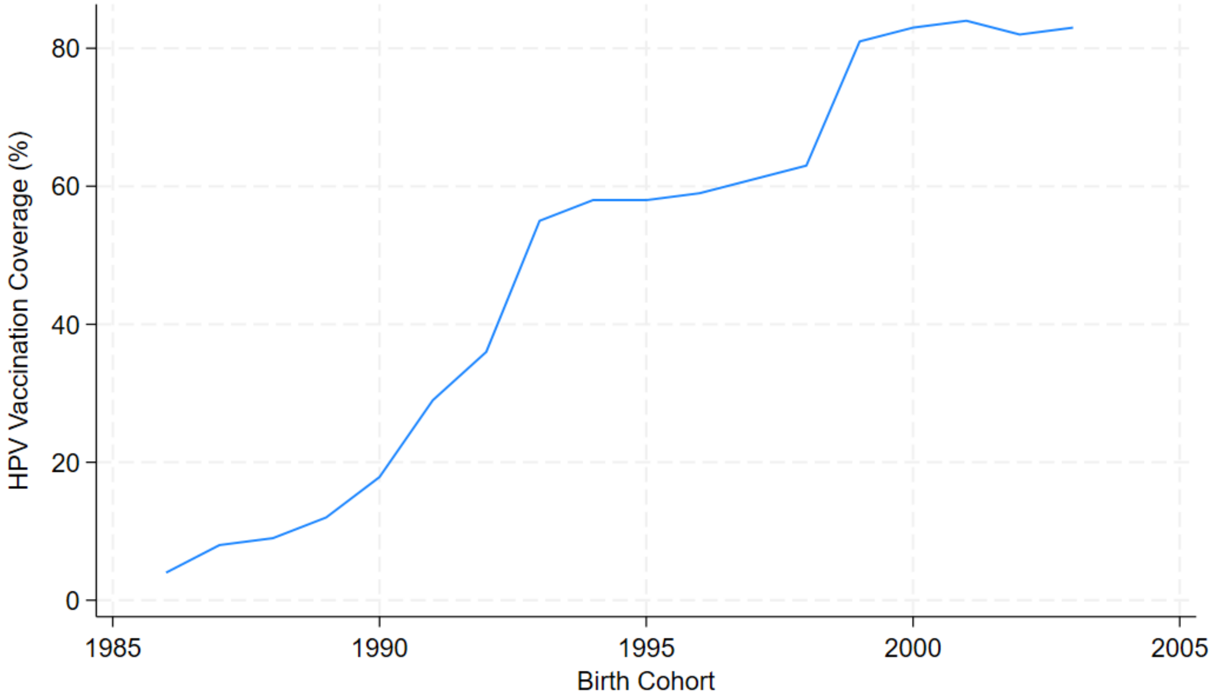


Figure S2: *Population-level vaccination coverage of at least one dose of a HPV vaccine in female birth cohorts 1986 to 2003. Vaccination coverage as reported by Public Health Agency of Sweden [1]*

Table S2: Age- and sex-stratified incidence rate ratio (IRR) of genital wart cases for post-vaccination time periods in reference to pre-vaccination (i.e. 2006-2007)

| **Age (years)** | **IRR (95%CI)** | | |
|---|---|---|---|
| **Women** | **2008-2011** | **2012-2015** | **2016-2018** |
| 15-19 | 0.90 (0.66 - 1.22) | 0.36 (0.25 - 0.50) | 0.11 (0.07 - 0.17) |
| 20-24 | 0.88 (0.81 - 0.95) | 0.51 (0.46 - 0.56) | 0.27 (0.25 - 0.29) |
| 25-29 | 0.88 (0.77 - 1.00) | 0.69 (0.60 - 0.79) | 0.50 (0.44 - 0.57) |
| 30-34 | 1.02 (0.92 - 1.14) | 0.90 (0.81 - 1.00) | 0.80 (0.72 - 0.90) |
| 35-39 | 1.16 (1.06 - 1.27) | 1.15 (1.05 - 1.25) | 1.14 (1.03 - 1.26) |
| 40-44 | 1.22 (1.12 - 1.33) | 1.18 (1.07 - 1.30) | 1.25 (1.13 - 1.38) |
| **Men** | | | |
| 15-19 | 1.19 (0.74 - 1.92) | 0.60 (0.36 - 0.99) | 0.21 (0.12 - 0.36) |
| 20-24 | 0.97 (0.89 - 1.05) | 0.65 (0.58 - 0.72) | 0.34 (0.30 - 0.40) |
| 25-29 | 1.02 (0.92 - 1.13) | 0.80 (0.72 - 0.88) | 0.60 (0.55 - 0.66) |
| 30-34 | 1.12 (0.98 - 1.29) | 1.03 (0.91 - 1.17) | 0.84 (0.74 - 0.95) |
| 35-39 | 1.19 (1.06 - 1.33) | 1.31 (1.18 - 1.46) | 1.19 (1.08 - 1.32) |
| 40-44 | 1.39 (1.22 - 1.59) | 1.42 (1.24 - 1.62) | 1.55 (1.36 - 1.78) |

**Section S1: Methods**

We obtained aggregated population counts through publicly available data from Statistics Sweden. The population data was aggregated by sex and categories of age and calendar year, separated by 1-unit intervals. They provided midyear population estimates used as time-at-risk in incidence rate (IR) calculations. Diagnoses of genital warts (GW) were obtained from the Prescribed Drug Register and Patient Register, and case counts were aggregated by 1-unit intervals of age, calendar year and cohort. The population data and GW case dataset were subsequently merged using Stata's *poprisktime* command, matching on unique values of age, period and cohort, and split by sex as a covariate [2].

In our analyses, we first calculated age-specific IR by period and birth cohort categories, and period-specific incidence rate by age groups. We illustrated these for men and women separately. Age groups were categorized as 15-19, 20-24, 25-29, 30-34, 35-39 and 40-44, and calendar periods were divided as 2006-2007, 2008-2011, 2012-2015 and 2016-2018. Meanwhile, birth cohorts were grouped as 1961-1985, 1986-1988, 1989-1992, 1993-1998 and 1999-2003. The crude IR were calculated by taking the total number of GW cases divided by person-years-at-risk in each corresponding category [3].

When examining the incidence changes by calendar period, we used Poisson models to estimate the incidence rate ratios (IRRs) with 95% confidence intervals (CIs) for each of the post-vaccination periods (i.e. 2008-2011, 2012-2015 and 2016-2018) using the pre-vaccination period, 2006-2007, as reference. We applied the Poisson regression model for each of the age groups and for females and males separately. We used a log-link function in the Poisson model with log person-time included as an offset, and robust standard errors to mitigate against model misspecification and variance heterogeneity.

Additionally, we also estimated the IRR of GW by comparing the incidence in birth cohorts categories eligible for different vaccination programs (i.e. 1989-1992, 1993-1998 and 1999-2003) using the youngest pre-vaccination cohort, 1986-1988, as reference . We used Poisson regression models with robust standard errors and included covariates of age and period as restricted cubic splines with 6 and 3 knots, respectively. Incidence of GW by birth cohorts were modelled separately for females and males.

Following the Poisson models comparing incidence by birth cohorts, we estimated the number of cases averted in each of these post-vaccination cohorts using the *margins* post-estimation command [4]. This was done by estimating the number of GW cases in each vaccinated birth cohort using each cohort's different estimated effects and size. This was compared with the corresponding expected number of GW cases when the cohort effect was forced to be the same as in the referent pre-vaccination cohort using the margin command in Stata [4]. The difference between estimated and expected GW cases was considered as the number of GW cases averted through HPV vaccination.

In the sensitivity analyses of the period analysis, we ran the same model including season as a covariate. Seasonality was included as a categorical variable with four strata representing spring (March-May), summer (June-August), autumn (September-November) and winter (December-February) based on the date of individual GW diagnoses or drug dispensation. In the cohort analyses, we used a negative binomial model to assess model sensitivity to the violation of the equidispersion assumption. The model included the independent variables in the same manner, and estimated both IRR and number of cases averted.

Table S3: Age- and sex-stratified incidence rate ratio (IRR) of genital wart cases for post-vaccination time periods in reference to pre-vaccination (i.e. 2006-2007), adjusted for seasonality

| **Age (years)** | **IRR (95%CI)** | | |
|---|---|---|---|
| **Women** | **2008-2011** | **2012-2015** | **2016-2018** |
| 15-19 | 0.90 (0.73 - 1.11) | 0.38 (0.30 - 0.47) | 0.13 (0.10 - 0.17) |
| 20-24 | 0.88 (0.77 - 1.00) | 0.51 (0.45 - 0.58) | 0.27 (0.23 - 0.31) |
| 25-29 | 0.88 (0.76 - 1.01) | 0.69 (0.60 - 0.79) | 0.50 (0.44 - 0.58) |
| 30-34 | 1.02 (0.90 - 1.16) | 0.90 (0.79 - 1.03) | 0.80 (0.70 - 0.92) |
| 35-39 | 1.16 (1.01 - 1.33) | 1.15 (1.00 - 1.31) | 1.14 (0.99 - 1.31) |
| 40-44 | 1.22 (1.06 - 1.41) | 1.18 (1.02 - 1.36) | 1.25 (1.07 - 1.46) |
| **Men** | | | |
| 15-19 | 1.16 (0.87 - 1.54) | 0.65 (0.48 - 0.88) | 0.28 (0.20 - 0.38) |
| 20-24 | 0.97 (0.85 - 1.10) | 0.65 (0.57 - 0.74) | 0.34 (0.30 - 0.40) |
| 25-29 | 1.02 (0.90 - 1.17) | 0.80 (0.70 - 0.91) | 0.60 (0.53 - 0.69) |
| 30-34 | 1.12 (0.98 - 1.29) | 1.03 (0.90 - 1.18) | 0.84 (0.73 - 0.97) |
| 35-39 | 1.19 (1.05 - 1.35) | 1.31 (1.15 - 1.49) | 1.19 (1.04 - 1.37) |
| 40-44 | 1.37 (1.18 - 1.60) | 1.40 (1.20 - 1.64) | 1.53 (1.30 - 1.80) |

Table S4: Incidence Rate Ratio (IRR) of genital warts and estimated number of cases averted by birth cohort

| **Birth Cohort** | **Vaccination Program(s)** | **IRR (95% CI)** | **Cases (95% CI)** |
|---|---|---|---|
| | **Women** | | |
| 1961-1985 | Pre-vaccination | 1.09 (1.01 - 1.16) | - |
| 1989-1992 | Subsidised | 0.75 (0.70 - 0.81) | 5,847 (4,347 - 7,347) |
| 1993-1998 | Catch-up | 0.36 (0.33 - 0.39) | 11,914 (10,523 - 13,304) |
| 1999-2003 | School-based | 0.07 (0.05 - 0.08) | 2,849 (2,511 - 3,188) |
| | **Men** | | |
| 1961-1985 | n/a | 1.19 (1.12 - 1.26) | - |
| 1989-1992 | n/a | 0.80 (0.75 - 0.84) | 4,788 (3,584 - 5,992) |
| 1993-1998 | n/a | 0.41 (0.38 - 0.45) | 8,073 (7,253 - 8,894) |
| 1999-2003 | n/a | 0.15 (0.12 - 0.18) | 853 (758 - 949) |

* Reference corresponds to cohort 1986-1988.

* Estimates modelled using negative binomial as part of **sensitivity analysis**

**Women**

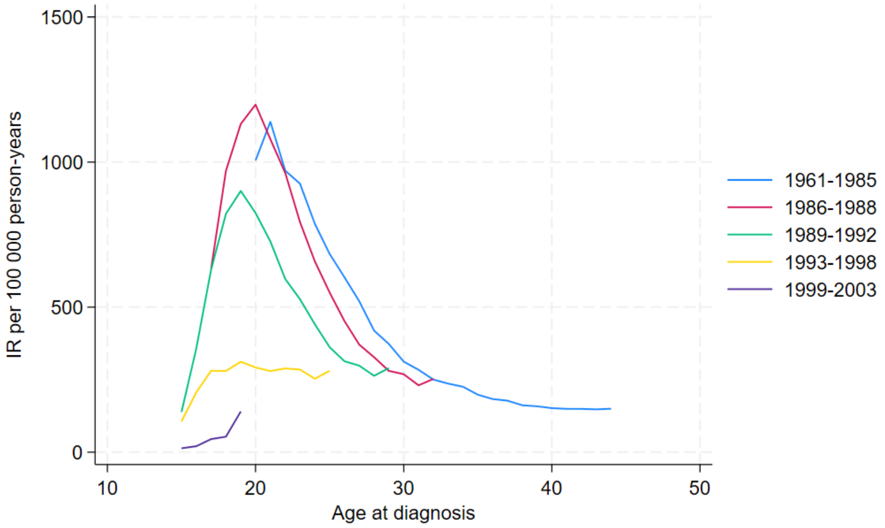


**Men**

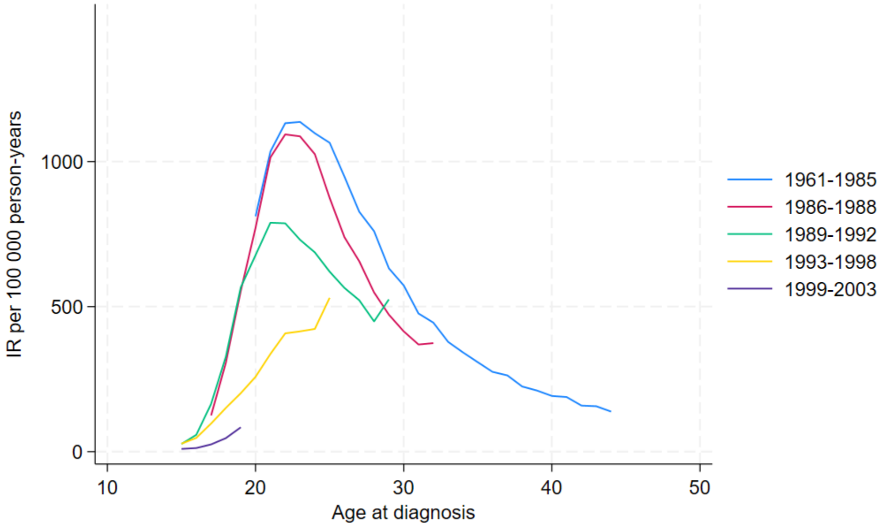


Figure S3: *Age-trend of genital warts incidence rates among women and men, stratified by cohort*
